# Leveraging Machine Learning and Clinical Data to Predict Response to Intralesional Corticosteroids in Keloid Patients

**DOI:** 10.1101/2025.05.05.25326913

**Authors:** Nina Zamani, Parand Akbari, Masoud Zamani, Cesar Antonio Rodriguez, Michael H. Tirgan, Akash Gunjan

**Author notes:** **Corresponding Author/Reprint Requests:** Akash Gunjan, PhD; 1115 West Call Street, Tallahassee, FL, 32306. **Funding Sources:** None. **IRB status:** The self-reported survey data used in this study was collected following IRB approval by the St. Luke’s-Roosevelt Hospital in New York. The study was subsequently transferred to Western IRB.

## Abstract

**Background:** Intralesional corticosteroid injections (ILCS) are a common treatment for keloid lesions; however, many patients exhibit resistance, and some experience worsening of their keloids following treatment.

**Objective:** To develop a machine learning (ML) tool capable of identifying factors that predict response to ILCS.

**Methods:** A keloid-specific survey database was accessed in May 2024. Various clinical and demographic factors were analyzed for correlation with self-reported responses to ILCS among 940 patients. Multiple ML models, including Neural Networks (NN) and Random Forest (RF), were trained on a subset of the survey data (training set) and tested on a separate subset (test set) to assess predictive accuracy.

**Results:** MM and RF models identified gender, keloid shape and age of the patients as the strongest determinants of ILCS response in keloid patients, achieving ∼95% predictive accuracy on our test dataset.

**Limitations:** The ML models were trained on self-reported survey data rather than data collected by clinicians, which may impact accuracy and reliability.

**Conclusions:** NN and RF based ML models using basic keloid patient data can be used to predict response to intralesional steroids in keloid patients accurately using a web-based user interface. Similar ML models maybe useful in clinical decision making regarding the use of steroid therapy for additional conditions.

**Capsule Summary:**

- Methods to predict response to intralesional corticosteroid treatment for keloids are unavailable but urgently needed given the significant number of patients who are refractory to steroid treatment.
- Machine learning algorithms based of self-reported keloid parameters and demographic information can be used to predict response to steroids prior to initiating therapy.

## INTRODUCTION

Keloid disorders are non-lethal fibroproliferative skin tumors/lesions characterized by excessive fibroblast proliferation and overproduction of extracellular matrix, particularly type I collagen [1]. Lesions extend beyond original wound boundaries and often cause pain, itching, and significant cosmetic and psychosocial distress. Management remains challenging due to high recurrence rates [2]. Intralesional corticosteroid injections (ILCS) is a mainline therapy for keloids, and results in dose-dependent reductions in inflammation, fibroblast activity, and collagen synthesis [3, 4]. Since its first reported efficacy in 1960 [4] ILCS remains widely used, with ongoing studies supporting its role in keloid regression [3].

Despite the widespread use of ILCS for keloids, patient response is inconsistent, with only about two-thirds of the patients benefiting to different extents, while a significant fraction experiences a worsening of symptoms [5-7]. Steroid resistance is observed in other conditions, including pediatric nephrotic syndrome [8], asthma [10], and inflammatory bowel diseases [9], highlighting variability in treatment response and the need for predictive tools.

Machine learning (ML) offers promising solutions in personalized medicine, enabling prediction of treatment outcomes by analyzing complex clinical datasets. ML has been successfully applied in diagnostics and treatment outcome prediction, enabling more tailored interventions [10]. By analyzing patient data, including demographic, genetic, and clinical information, ML models can identify patterns that may not be apparent through traditional analysis [11]. In oncology, algorithms like random forests (RF) and neural networks (NN) predict survival outcomes [12], while in surgery, ML models have optimized treatment planning and improved patient safety [13].

Here we have applied ML tools to predict steroid treatment outcomes based on self-reported keloid patient survey data. By identifying patients unlikely to benefit from ILCS, clinicians can recommend alternative therapies, improving outcomes and reducing ineffective treatments.

## METHODS

Please see the Supplementary Information for a detailed description of the dataset and the ML tools used in this study.

Our workflow for the study is shown in Figure 1A. The ML framework was applied on an updated version of the IRB approved, self-reported online keloid patient survey data that was published previously [6], and has been updated here in Supplementary Figures S1-S4. The dataset included a total of 1,940 keloid patients from whom 940 patients (290 male, 650 female) provided sufficient information on their steroid treatment to be included in our analysis. The dataset included demographic details, keloid parameters and history, treatment responses, and triggering factors. These features are summarized in Figure 1b. After preprocessing, numerical features were standardized, categorical variables were one-hot encoded, and class imbalance was addressed using SMOTE-ENN. We applied multiple ML models, including RF, Gradient Boosting (GB), XGBoost, NN, and Support Vector Classifier (SVC), with hyperparameter tuning and 5-fold cross-validation for performance evaluation. Feature importance was analyzed using Shapley Additive Explanations (SHAP) and permutation importance to enhance interpretability. Performance metrics included accuracy, precision, recall, F1-score, and Area Under the Curve (AUC) scores.

**Figure 1.**
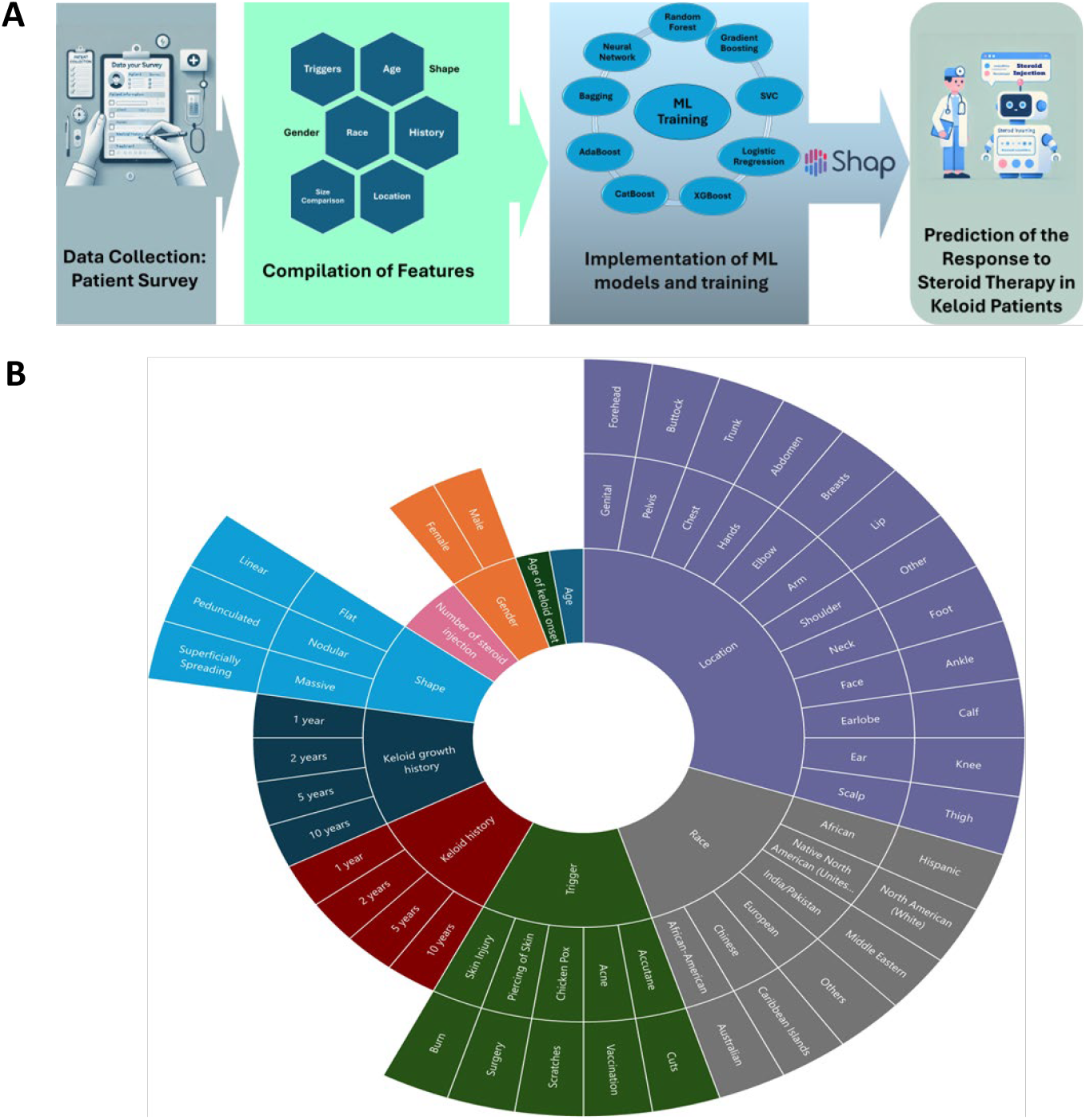
Overview of the workflow and clinical data features employed in this study. **(A)** Overall workflow of the study. **(B)** Features employed in the machine Learning (ML) models.

## RESULTS

In this study, the predictive capabilities of the ML models applied to the dataset are presented. A variety of models were trained and evaluated using a range of performance metrics, including accuracy, precision, recall, F1 score, Receiver Operating Characteristic Area Under the Curve (ROC-AUC), F0.5 score, and Precision-Recall Area Under the Curve (PR-AUC). Additionally, cross-validation techniques were employed to ensure reliable and robust results. Several visualization techniques, including heatmaps and spider plots, were used to offer a comparative view of model performance across all metrics.

### Performance of different ML models in predicting response to steroid therapy for keloids

The performance of various ML models is summarized in Table 1, listing accuracy, precision, recall, F1 score, ROC-AUC-OVR, F0.5 score, and PR-AUC. Figures 2A and 2B visually compare model performances via a heatmap and a spider plot. NN achieved the highest accuracy (0.949 ± 0.013), precision (0.951 ± 0.012), recall (0.949 ± 0.013), and F1 score (0.945 ± 0.015). Its ROC-AUC of 0.957 ± 0.041 further confirms strong class distinction. RF closely followed, achieving 0.947 ± 0.025 accuracy and a higher ROC-AUC of 0.987 ± 0.024. Support Vector Classifier (SVC) showed good accuracy (0.945 ± 0.016) but lower ROC-AUC (0.907 ± 0.052). Gradient Boosting (GB) and CatBoost performed well, with accuracies of 0.935 ± 0.024 and 0.929 ± 0.023 and ROC-AUC values of 0.976 ± 0.041 and 0.970 ± 0.030, respectively. XGBoost (accuracy 0.939 ± 0.030, ROC-AUC 0.962 ± 0.054) and Bagging (accuracy 0.907 ± 0.043) also showed solid performance. PR-AUC values remained consistently high across models, mostly above 0.95, confirming strong precision-recall balance. While NN showed slightly better precision, recall, and F1 scores, RF demonstrated comparable or superior ROC-AUC, indicating robust class-separation. In practice, NN may be preferred for scalability, while RF for easier interpretability and feature analysis.

**Table 1:** Performance metrics for various models sorted by accuracy

| Model | Accuracy | Precision | Recall | F1 Score | ROC-AUC-OVR | F0.5 Score | PR-AUC |
| --- | --- | --- | --- | --- | --- | --- | --- |
| Neural Network | 0.949±0.013 | 0.951±0.012 | 0.949±0.013 | 0.945±0.015 | 0.957 ± 0.041 | 0.947±0.014 | 0.974±0.007 |
| Random Forest | 0.947±0.025 | 0.951±0.023 | 0.947±0.025 | 0.944±0.027 | 0.987 ± 0.024 | 0.946±0.025 | 0.975±0.011 |
| SVC | 0.945±0.016 | 0.946±0.017 | 0.945±0.016 | 0.941±0.019 | 0.907 ± 0.052 | 0.942±0.019 | 0.972±0.008 |
| XGBoost | 0.939±0.030 | 0.944±0.027 | 0.939±0.030 | 0.936±0.029 | 0.962 ± 0.054 | 0.939±0.028 | 0.972±0.009 |
| Logistic Regression | 0.939±0.017 | 0.940±0.017 | 0.939±0.017 | 0.933±0.020 | 0.922 ± 0.035 | 0.935±0.019 | 0.969±0.008 |
| Gradient Boosting | 0.935±0.024 | 0.940±0.018 | 0.935±0.024 | 0.933±0.021 | 0.976 ± 0.041 | 0.936±0.019 | 0.972±0.004 |
| CatBoost | 0.929±0.023 | 0.932±0.022 | 0.929±0.023 | 0.922±0.026 | 0.970 ± 0.030 | 0.925±0.025 | 0.966±0.010 |
| Bagging | 0.907±0.043 | 0.918±0.029 | 0.907±0.043 | 0.901±0.037 | 0.963 ± 0.064 | 0.907±0.033 | 0.961±0.009 |
| AdaBoost | 0.907±0.067 | 0.928±0.031 | 0.907±0.067 | 0.909±0.055 | 0.934 ± 0.041 | 0.918±0.041 | 0.969±0.007 |

**Figure 2.**
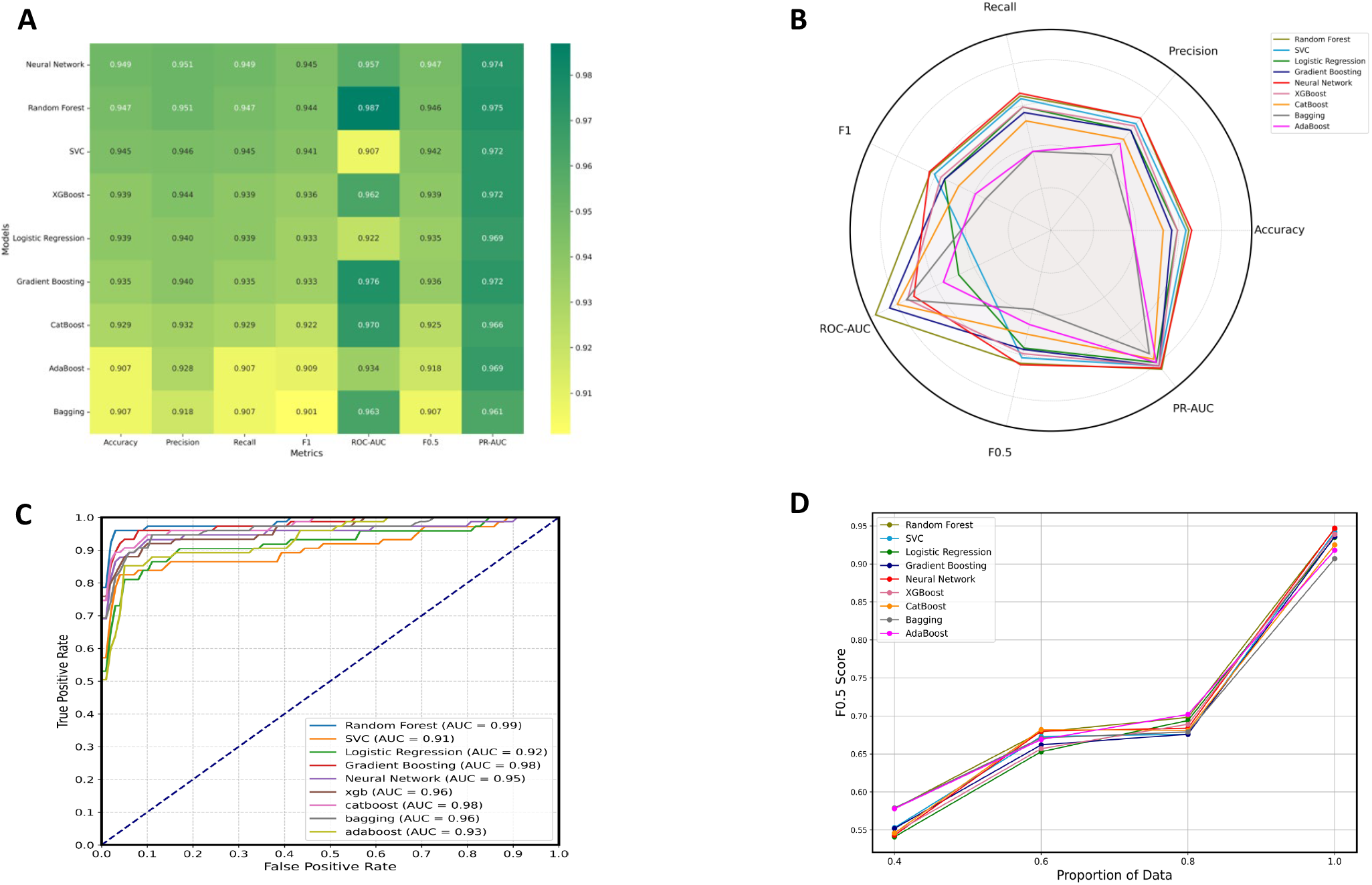
(A) Heatmap of performance metrics for various ML models, sorted by accuracy. **(B)** Spider plot illustrating the performance metrics for various ML models. **(C)** AUC values for the different ML models. **(D)** Effect of training set size on F0.5 scores across different ML models. Increasing the proportion of data significantly enhances performance, underscoring the positive impact of larger training sets on model effectiveness.

### ROC-AUC comparison of different ML models

The performance of the ML models was evaluated using ROC curves, which are shown in Figure 2C. Each model is represented by a distinct line on the plot, illustrating the relationship between the true positive rate (sensitivity) and the false positive rate (1−specificity) across different classification thresholds. A model that performs better will have a curve that is closer to the top-left corner, indicating higher sensitivity with fewer false positives. The AUC was calculated for each model to quantify performance, with higher AUC values signifying more effective models.

The AUC values for the models are as follows: RF (0.99), SVC (0.91), Logistic Regression (0.92), GB (0.98), NN (0.95), XGBoost (0.96), Cat-Boost (0.98), Bagging (0.96), and AdaBoost (0.93). Among these, RF showed the highest AUC scores, indicating excellent classification performance with near-perfect sensitivity and specificity. All models exhibit strong performance, with AUC values above 0.90, reflecting their effectiveness in accurately classifying the dataset.

### Effect of training set size on model accuracy

The impact of training set size on regression accuracy was assessed by randomly selecting 40%, 60%, 80%, and 100% of the training data, followed by 5-fold cross-validation across ML models. Figure 2D shows that F0.5 scores improve with larger training sets, indicating higher prediction accuracy.

### Model interpretability and importance of individual features

Feature importance was analyzed using SHAP and permutation methods to identify key clinical and demographic factors influencing model predictions and steroid therapy outcomes in keloid patients.

#### SHAP Results

The global SHAP bar plot (Figure 3A) shows the mean absolute SHAP values for each feature group, highlighting patient gender and keloid morphology as the strongest predictors of steroid therapy outcomes. Female patients were more likely to be predicted as sensitive to treatment, while males tended toward resistance. Similarly, lesion morphology—flat, nodular, or massive—strongly influenced predictions, with flat and nodular lesions associated with sensitivity and massive lesions with resistance. Growth history also had a major impact: patients whose keloids continued growing over 1, 2, 5, or 10 years were more likely to be predicted as resistant, while other factors like number of steroid injections, age of onset, and lesion location contributed moderately. Initial models included growth history, intuitively a strong predictor since persistent growth after multiple steroid injections often signals resistance. However, because clinical scenarios often involve treatment-naïve patients or missing records, feature ablation analysis was performed. Removing growth history, number of injections, or both led to a modest drop in model accuracy (from ∼95% to ∼90%), demonstrating that models remained clinically valuable without these features. The SHAP summary plot (Figure 3B) provided patient-level insights, with each dot representing how a feature shifted predictions toward sensitivity (positive SHAP value) or resistance (negative SHAP value). Red dots indicated higher feature values (e.g., female gender), blue lower values (e.g., male gender). Analysis showed that flat or nodular morphology and female gender favored sensitivity, while massive morphology and growth history favored resistance. Finally, SHAP interaction analysis (Figure 3C) explored joint effects of age and gender. Younger female patients showed strong positive interaction effects (greater likelihood of steroid sensitivity), but this reversed with aging, where older females trended toward resistance. In contrast, younger males were more resistant, but older males showed interaction values moving toward zero or slightly positive, suggesting reduced resistance with age. These interactions highlight that feature relationships evolve with patient demographics, underscoring the importance of analyzing feature combinations rather than isolated effects.

**Figure 3.**
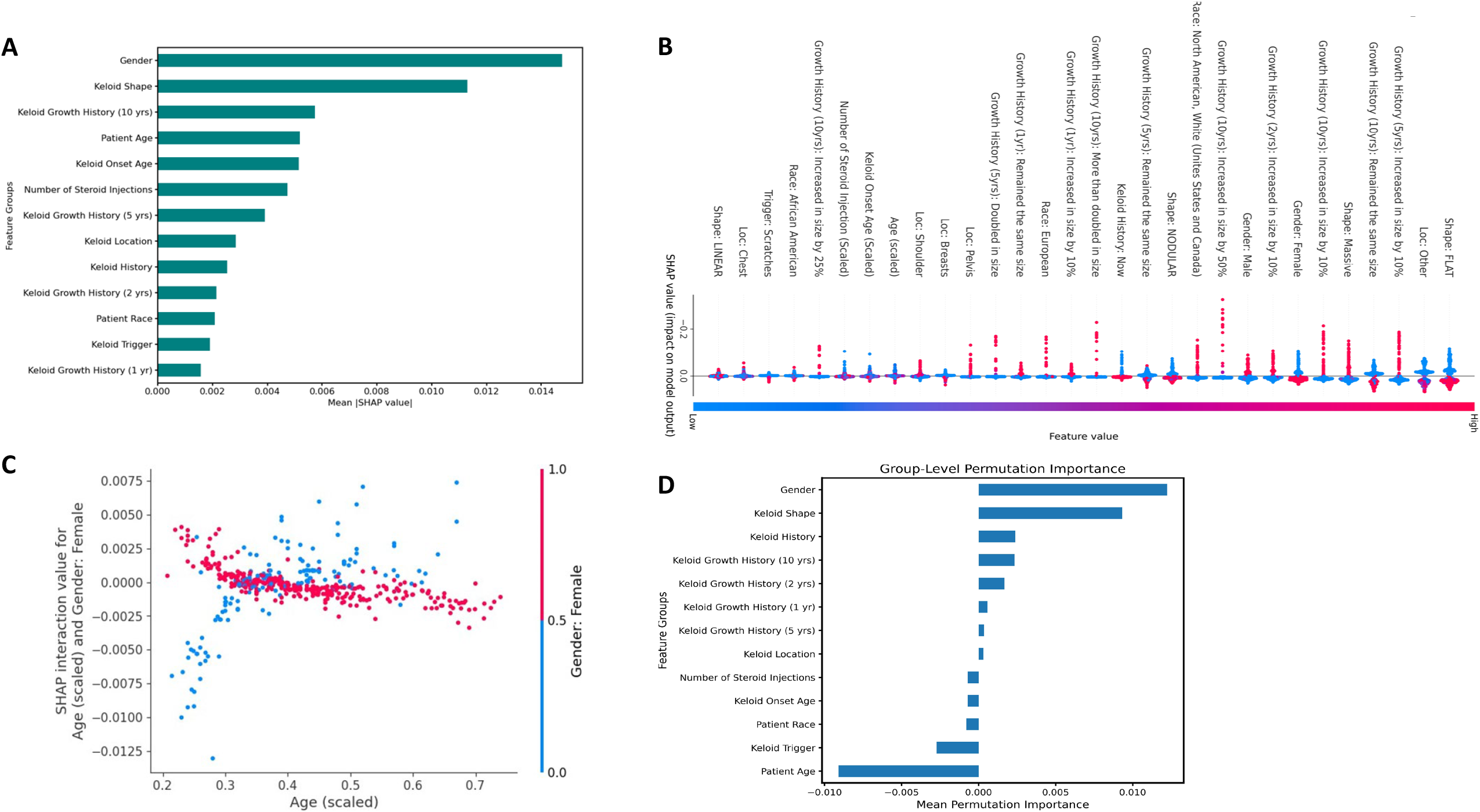
SHAP parameters showing the relative importance of different features, as well as interactions between specific features in the model. **(A)** Global SHAP bar plot illustrating the average importance of feature groups in determining the model’s prediction of steroid therapy sensitivity. Patient gender and the morphology of keloid lesions exhibit the highest mean SHAP values, highlighting their strong influence on treatment outcomes. **(B)** SHAP summary plot showing the effect of individual features on the predicted response to steroid therapy. Positive SHAP values indicate sensitivity to treatment, while negative values suggest resistance. The color gradient represents feature values, where red indicates high values and blue indicates low values. Notably, patient gender and the morphology of keloid lesions exhibit a range of effects depending on the specific patient. **(C)** SHAP Interaction Plot revealing the interaction between age and gender in predicting response to steroid therapy. Positive SHAP interaction values indicate that the interaction drives the prediction toward sensitivity, while negative values suggest resistance. This plot suggests that younger female patients are more likely to be sensitive to treatment, with a diminishing effect as age increases. **(D)** Group-level permutation importance plot highlighting the standalone effect of feature groups on model accuracy. Patient gender and the morphology of keloid lesions remain the most critical predictors, reinforcing findings from the SHAP analysis, though age demonstrates less direct influence.

#### Permutation Importance

To complement SHAP analysis, we applied permutation importance to assess each feature’s standalone contribution by measuring any reduction in model performance when feature values were shuffled. The group-level permutation plot (Figure 3D) confirmed that patient gender and keloid morphology are the most important factors individually. While age showed strong influence in SHAP analysis through interaction with gender, permutation results revealed low standalone importance, suggesting age impacts predictions mainly through interactions. Together, these analyses provide a comprehensive understanding of how clinical and demographic factors influence model predictions, aiding in personalizing steroid therapy for keloids.

### Web-based tool for clinicians to predict steroid response

We developed a simple web-based tool for clinicians to predict steroid response for keloids at the point of care (Figure 4). Clinicians can input the relevant data to obtain a prediction regarding the use of steroids for keloid therapy in individual patients.

**Figure 4.**
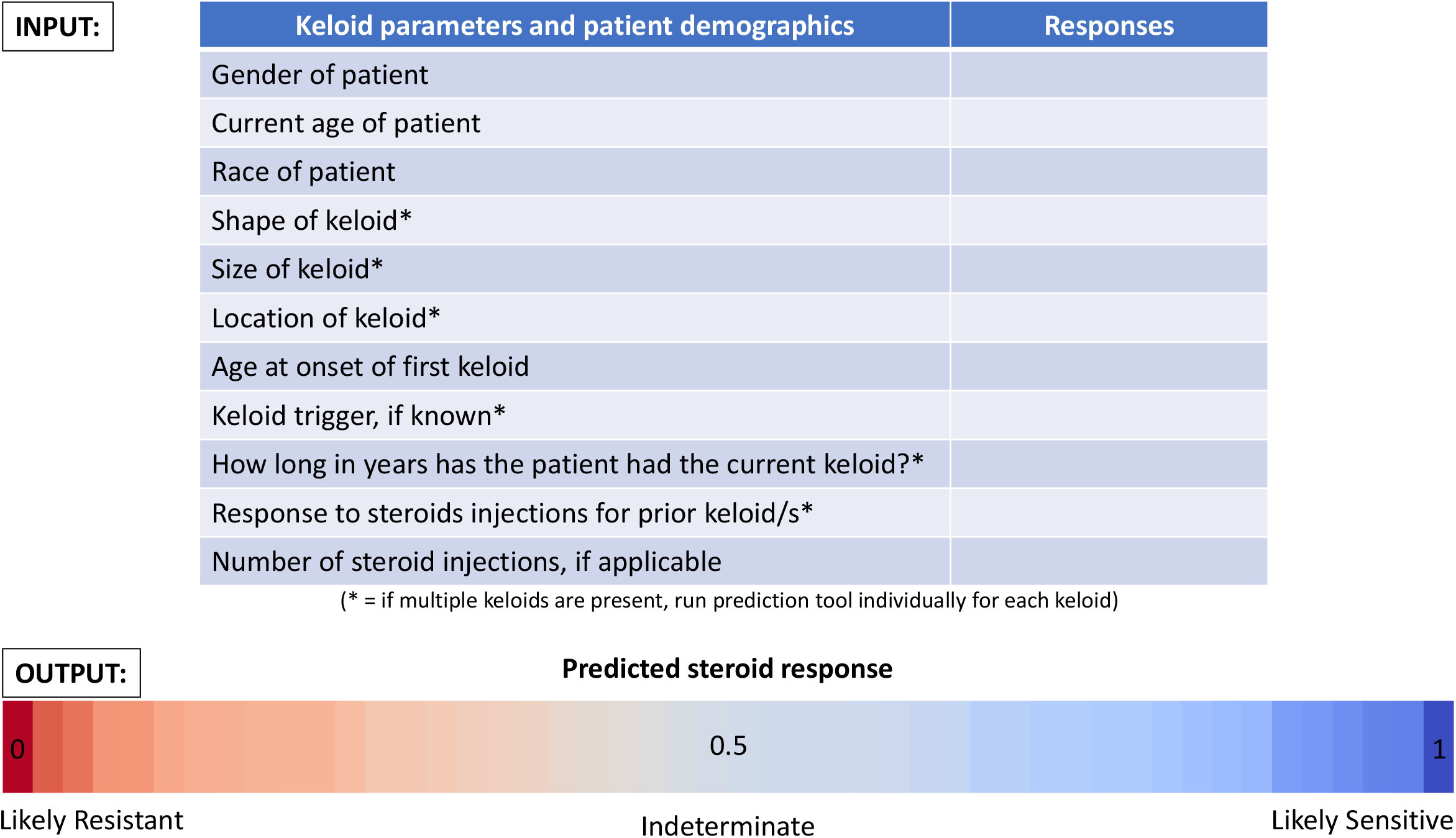
A web-based tool employing the NN model with a simple user interface for clinicians to predict steroid response in keloid patients. Clinicians would simply input the relevant keloid parameters and patient demographic information to obtain a prediction on the potential steroid response for individual keloid patients to guide their clinical decision making.

## DISCUSSION

We evaluated nine ML models to predict keloid patient responses to corticosteroid therapy, assessing performance across seven metrics. NN and RF achieved top-tier results, with NN showing slightly higher accuracy and F1 score, while RF demonstrated marginally higher ROC-AUC. Both models offer strong predictive power; however, NN was selected as the primary model for its consistent performance and scalability. Clinical and demographic data analysis identified gender as the strongest predictor, with female patients more likely to respond favorably to therapy. Notably, women under 45 showed higher sensitivity to ILCS therapy, whereas older women exhibited resistance, potentially reflecting hormonal changes post-menopause. These findings highlight the importance of gender-specific factors in predicting steroid treatment outcomes for keloid disorder.

Our findings are consistent with published studies suggesting a gender bias in response to steroid therapy. Several studies have reported that females respond less favorably to corticosteroid treatments compared to males [14-17]. For example, older female patients with inflammatory bowel disease (IBD) are more likely to exhibit prednisone resistance or dependence compared to younger females and males [14]. This gender disparity in steroid response may be underpinned by the sexually dimorphic effects of glucocorticoids. Crosstalk between glucocorticoid and sex steroid hormones significantly impacts key processes like metabolism and immune regulation [15]. For instance, glucocorticoid receptor (GR) signaling can be modulated by estrogen receptor (ERα and ERβ) pathways, suggesting a biological basis for the observed differences in therapeutic outcomes between males and females [16]. Furthermore, in systemic lupus erythematosus (SLE), a disease with a striking female predominance (9:1 female-to-male ratio), estradiol-mediated activation of ERα has been implicated in disease progression. Blocking ERα action with antagonists such as Fulvestrant has been shown to improve disease indicators, pointing to the significance of steroid receptor crosstalk in disease mechanisms [17]. These insights into the interplay between GR and ERα pathways underscore the need to consider gender as a critical factor in optimizing steroid-based therapies. Addressing these differences is essential for improving outcomes and tailoring treatments for conditions with significant gender disparities in steroid response.

The morphology of keloid lesions emerged as the second most important feature in predicting steroid sensitivity, with flat and nodular keloid lesions showing greater responsiveness compared to massive keloid lesions. While a previous study focused on keloid lesion contour as a predictor of steroid response, finding that patients with higher contour scores were more likely to respond to corticosteroid injections within a 3-month follow-up period [18], our findings may not necessarily contradict this. Contour and morphology are related parameters, with higher contour (in the Z-plane) potentially correlating with smaller keloid lesions in the X-Y plane, leading to a more nodular appearance. Our study, however, utilized self-reported patient data collected over a longer timeframe, capturing the long-term impact of keloid behavior and response to steroid treatment. This broader perspective may provide a more comprehensive understanding of the relationship between keloid morphology and steroid sensitivity.

The growth history of keloids was the third most important feature in predicting response to steroids, indicating that patients with ongoing keloid growth for 1 to 10 years were more likely to exhibit steroid resistance. This finding is consistent with data from online keloid disorder management resources, which similarly highlight that persistent growth over extended periods is associated with poor response to steroid therapy [19]. Removing the growth history of keloids from our models resulted in little or no decrease in the predictive values of our models.

Age is also one of the important factors in steroid sensitivity in keloid patients. This finding aligns with existing evidence suggesting that age plays a significant role in corticosteroid response across various medical conditions. In general, younger patients often exhibit more robust responses to steroids compared to older individuals, which correlates with the duration of the illness, and age of the keloids. This phenomenon may be attributed to age-related changes in immune function, metabolism, and tissue repair mechanisms, which can influence how steroids exert their anti-inflammatory and immunosuppressive effects. For asthma and autoimmune diseases, studies have reported decreased steroid efficacy in older patients [8, 19-21], potentially due to age-related reductions in glucocorticoid receptor sensitivity or altered inflammatory pathways. The observed relationship between age and steroid sensitivity in keloid patients may similarly reflect underlying biological changes that affect fibroblast proliferation, collagen synthesis, and overall wound healing processes. Currently, there are no clinical data on the relationship between age and steroid resistance in keloid patients. Future clinical studies should address this gap, as understanding age-related differences is crucial for clinicians to make informed decisions about selecting the most appropriate therapies.

The age of onset and location of keloids, as well as triggering factors were also significant predictors of steroid sensitivity. Earlier onset of keloid lesions correlated with varying levels of treatment response, suggesting that the age at which keloid lesions first develop may influence their sensitivity to steroid therapy. The initial trigger for the formation of keloid lesions, such as acne, surgery, or injury, highlighted by SHAP analysis, also played a role in predicting outcomes, with different triggers potentially affecting keloid behavior and response to treatment. Furthermore, the anatomical location of the keloid influenced responsiveness, as certain locations were more likely to show favorable outcomes on steroid therapy compared to others. These findings emphasize the multifactorial nature of keloid disorder and the importance of considering these clinical variables when developing personalized therapies.

In conclusion, this study demonstrates the potential of ML models to improve the prediction of response to intralesional steroids in keloid patients through a simple web-based interface. By identifying key predictors such as gender, keloid lesion morphology, growth history, and other clinical factors, we provide insights that can guide more effective and personalized treatment strategies. These findings underscore the importance of considering patient-specific characteristics in clinical practice and highlight the need for further research to validate these results and explore the underlying biological mechanisms. Such advancements will help optimize therapeutic decisions and improve outcomes for keloid patients. Finally, although our current models were developed using a large dataset specifically from keloid patients, it is likely that similar approaches would yield useful ML models for predicting response to steroid therapy for other conditions as well.

## Supporting information

Supplemental info including detailed methods

## Data Availability

All data produced in the present study are available upon reasonable request to the authors

## Abbreviations

(AUC): Area Under the Curve
(AI): Artificial Intelligence
(GB): Gradient Boosting
(ILCS): Intralesional Corticosteroid
(ML): Machine Learning
(NN): Neural Networks
(PR-AUC): Precision-Recall Area Under the Curve
(RF): Random Forrest
(ROC-AUC): Receiver Operating Characteristic Area Under the Curve
(SHAP): Shapley Additive Explanations
(SMOTE-ENN): Over-sampling using SMOTE and cleaning using ENN
(SVC): Support Vector Classifier
(eXtreme Gradient Boosting): XGBoost
GB: (Gradient Boosting)
(Categorical Boosting): CatBoost
SVC: (Support Vector Classifier)

## REFERENCES

1. Zhang, X., X. Wu, and D. Li, The Communication from Immune Cells to the Fibroblasts in Keloids: Implications for Immunotherapy. International Journal of Molecular Sciences, 2023. 24(20): p. 15475.

2. Chike-Obi, C.J., P.D. Cole, and A.E. Brissett, Keloids: pathogenesis, clinical features, and management. 2009. p. 178--184.

3. Atiyeh, B.S., Nonsurgical management of hypertrophic scars: evidence-based therapies, standard practices, and emerging methods. Aesthetic plastic surgery, 2007. 31: p. 468--492.

4. Sexton, G.B., Local Injection of Triamcinolone Acetonide in the Management of Certain Skin Conditions:(Preliminary Report). Canadian Medical Association Journal, 1960. 83(26): p. 1379.

5. Tirgan, M. Worsening of keloids after intralesional injections. in Journal of the American Academy of Dermatology. 2013. MOSBY-ELSEVIER 360 PARK AVENUE SOUTH, NEW YORK, NY 10010–1710 USA.

6. Tirgan, M., Intralesional triamcinolone acetonide in the treatment of keloid lesions-can the treatment be harmful to some patients? results of an online survey. Int J Keloid Res, 2017. 1: p. 21–28.

7. Muneuchi, G., et al., Long-term outcome of intralesional injection of triamcinolone acetonide for the treatment of keloid scars in Asian patients. Scandinavian journal of plastic and reconstructive surgery and hand surgery, 2006. 40(2): p. 111--116.

8. Adcock, I.M., et al., Steroid resistance in asthma: mechanisms and treatment options. Current allergy and asthma reports, 2008. 8(2): p. 171–178.

9. Coates, E., et al., Patient preferences and current practice for adults with steroid-resistant ulcerative colitis: POPSTER mixed-methods study. Health Technology Assessment (Winchester, England), 2022. 26(41): p. 1.

10. Sidey-Gibbons, J.A.M. and C.J. Sidey-Gibbons, Machine learning in medicine: a practical introduction. BMC medical research methodology, 2019. 19: p. 1--18.

11. Pfob, A., S.-C. Lu, and C. Sidey-Gibbons, Machine learning in medicine: a practical introduction to techniques for data pre-processing, hyperparameter tuning, and model comparison. BMC medical research methodology, 2022. 22(1): p. 282.

12. Huang, Y., et al., Application of machine learning in predicting survival outcomes involving real-world data: a scoping review. BMC medical research methodology, 2023. 23(1): p. 268.

13. Arjmandnia, F. and E. Alimohammadi, The value of machine learning technology and artificial intelligence to enhance patient safety in spine surgery: a review. Patient Safety in Surgery, 2024. 18(1): p. 11.

14. Lucafò, M., et al., Gender may influence the immunosuppressive actions of prednisone in young patients with inflammatory bowel disease. Frontiers in Immunology, 2021. 12: p. 673068.

15. Cvoro, A., et al., Cross talk between glucocorticoid and estrogen receptors occurs at a subset of proinflammatory genes. The Journal of Immunology, 2011. 186(7): p. 4354–4360.

16. Rider, V., et al., Gender bias in human systemic lupus erythematosus: a problem of steroid receptor action? Frontiers in immunology, 2018. 9: p. 611.

17. Lee, I., et al., Gender differences in prednisone adverse effects: Survey result from the MG registry. Neurology: Neuroimmunology & Neuroinflammation, 2018. 5(6): p. e507.

18. Ud-Din, S., et al., Identification of steroid sensitive responders versus non-responders in the treatment of keloid disease. Archives of dermatological research, 2013. 305: p. 423–432.

19. Juckett, G. and H. Hartman-Adams, Management of keloids and hypertrophic scars. American family physician, 2009. 80(3): p. 253–260.

20. Jaiswal, A.K., et al., Short palate, lung, and nasal epithelial clone 1 (SPLUNC1) level determines steroid-resistant airway inflammation in aging. American Journal of Physiology-Lung Cellular and Molecular Physiology, 2022. 322(1): p. L102–L115.

21. Borkar, N.A., C.K. Combs, and V. Sathish, Sex steroids effects on asthma: a network perspective of immune and airway cells. Cells, 2022. 11(14): p. 2238.

