## Supplemental figures for "Leveraging Machine Learning and Clinical Data to Predict Response to Intralesional Corticosteroids in Keloid Patients"

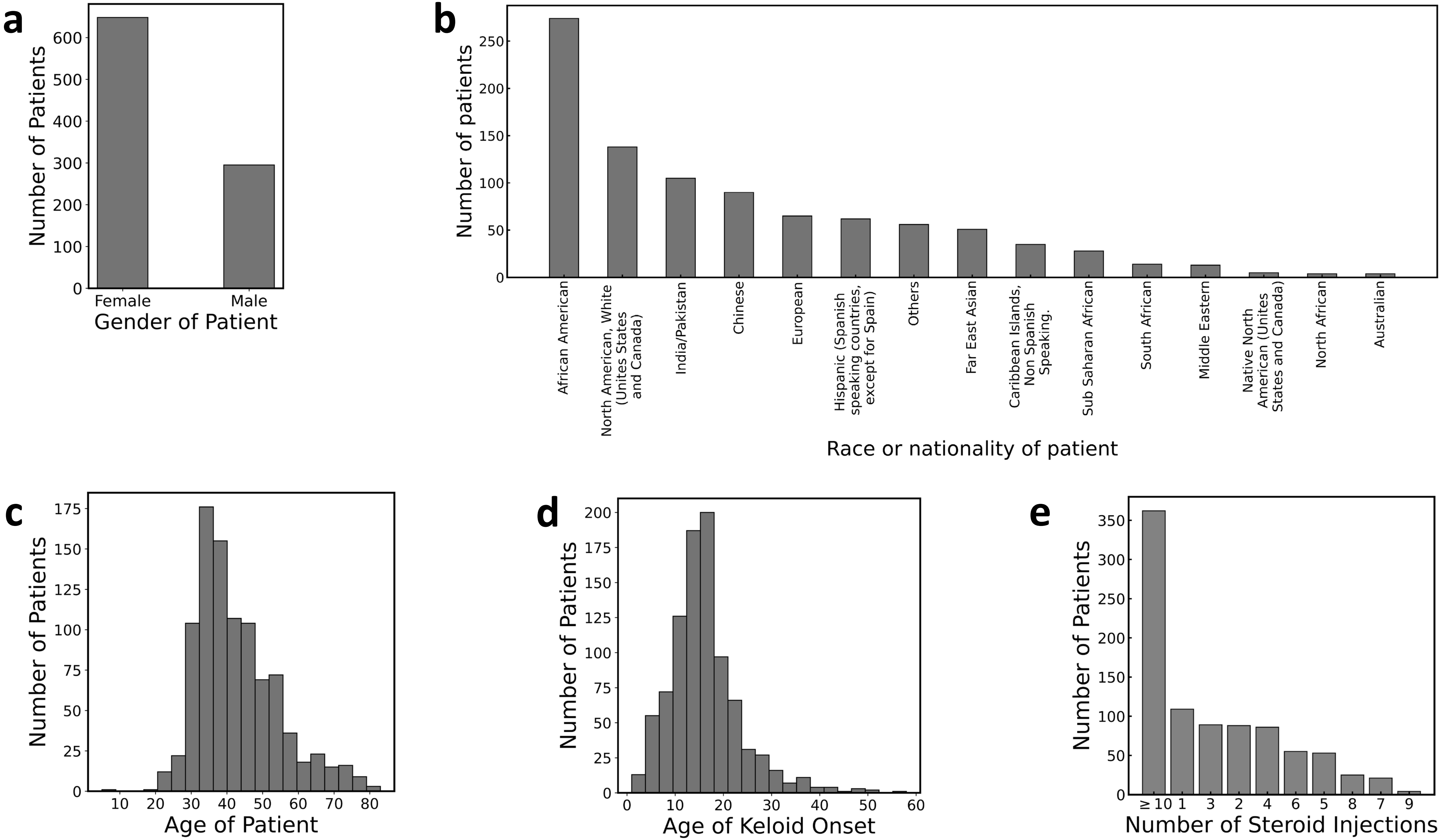


**Supplementary Figure S1**

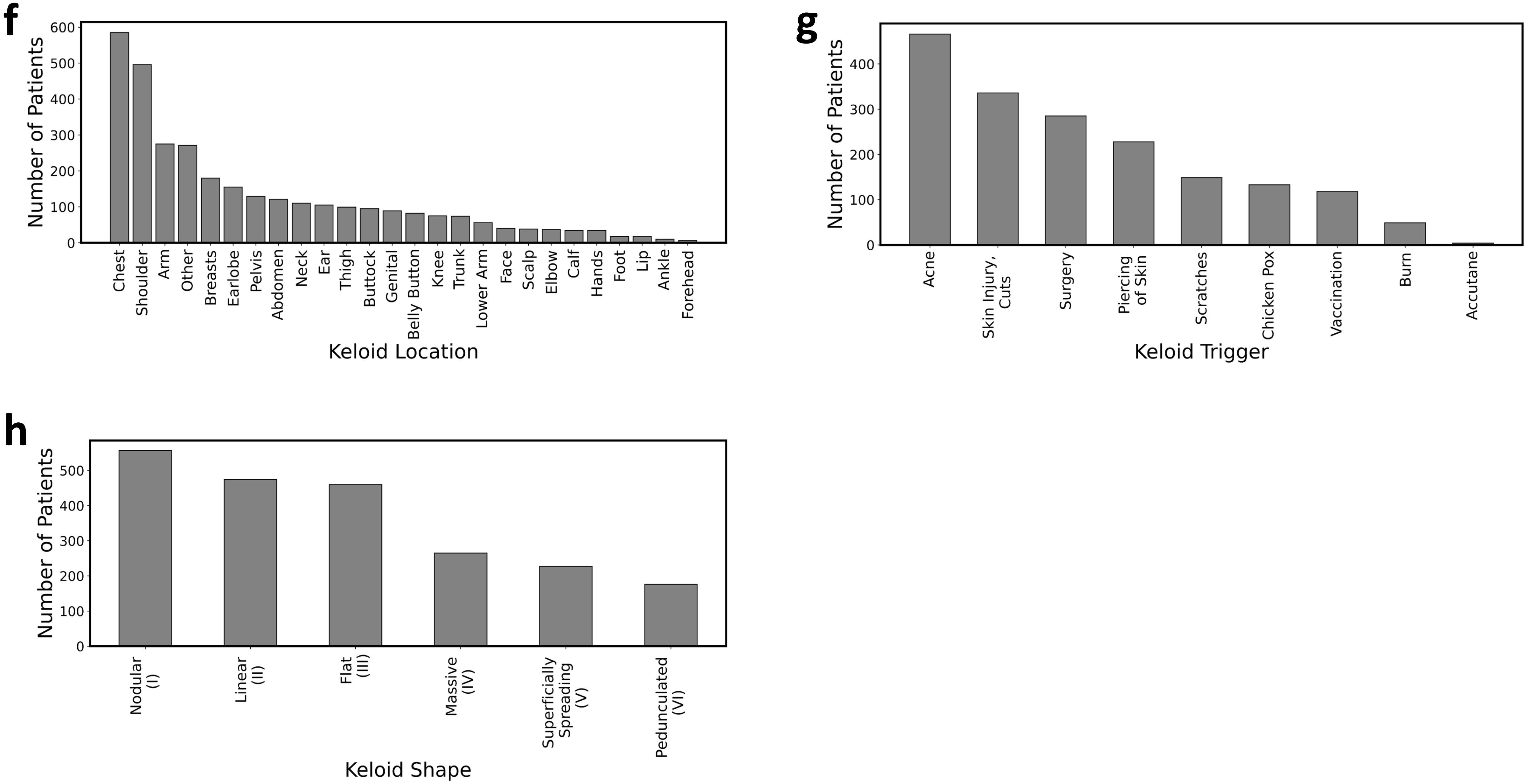


**Supplementary Figure S1**

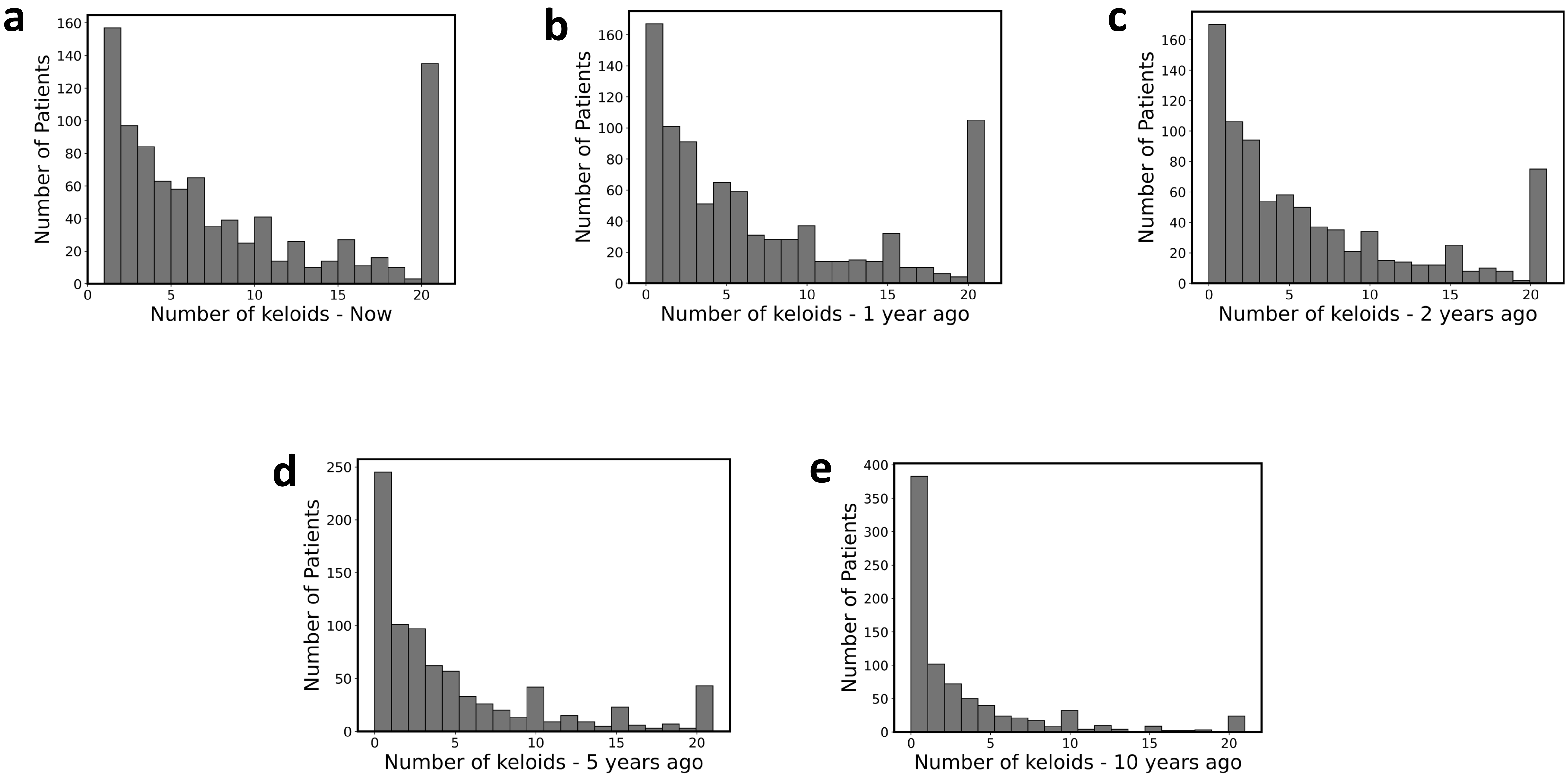


**Supplementary Figure S2**

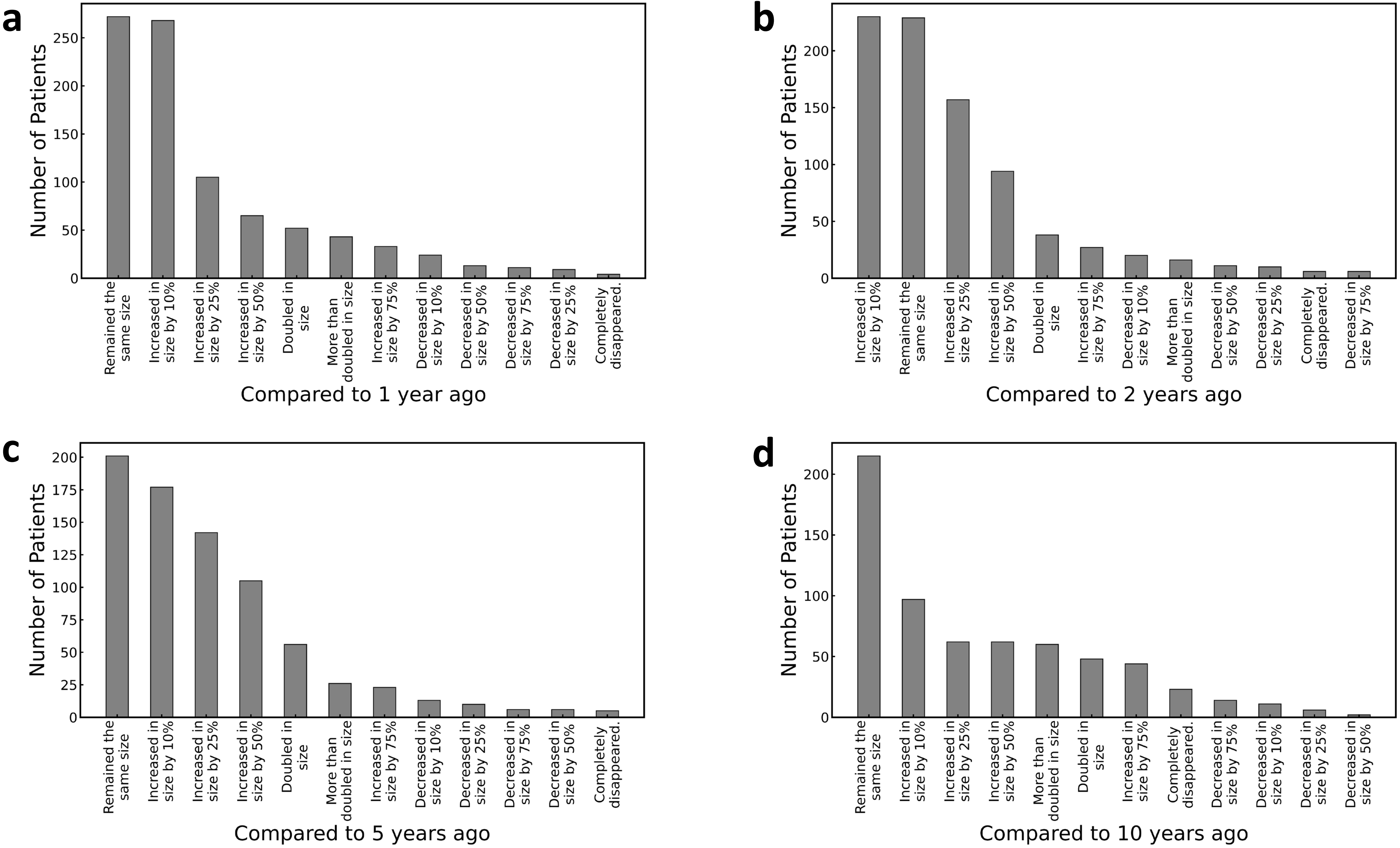


**Supplementary Figure S3**

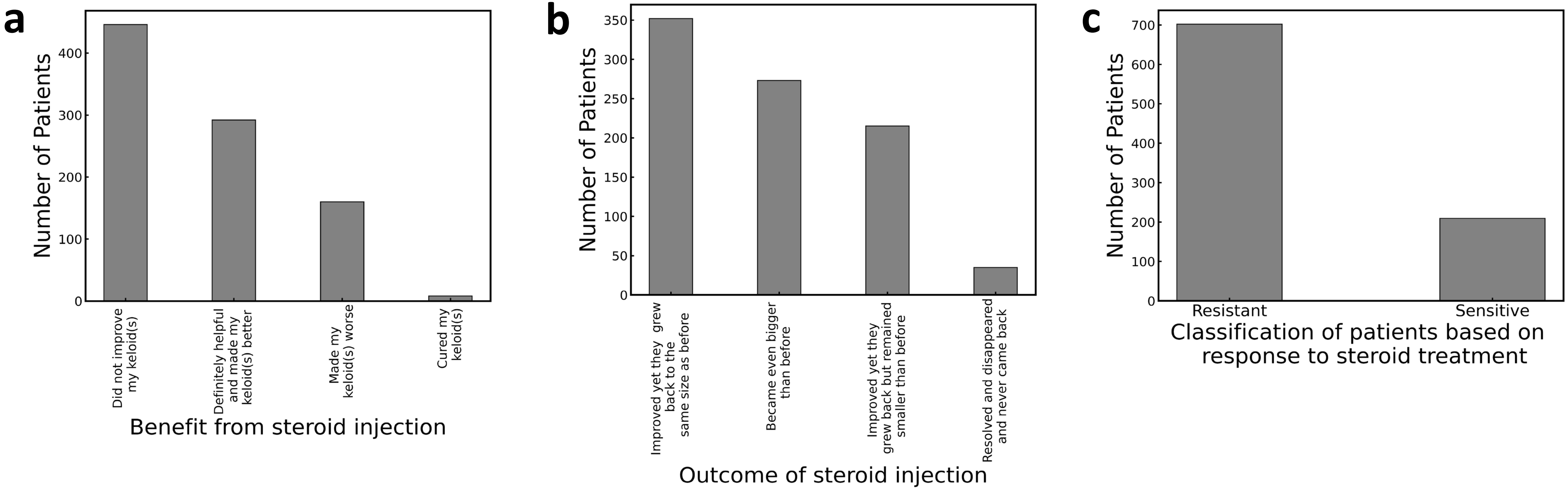


Supplementary Figure S4
