## Supplemental info including detailed methods for "Leveraging Machine Learning and Clinical Data to Predict Response to Intralesional Corticosteroids in Keloid Patients"

**SUPPLEMENTARY INFORMATION**

### METHODS

Details of the processes involved in gathering the dataset, engineering features, and the machine learning (ML) algorithms used are provided below.

**Dataset:** A comprehensive questionnaire was designed to survey a large, comprehensive cohort of patients diagnosed with keloid disorder. Initially approved by the Institutional Review Board (IRB) of St. Luke's-Roosevelt Hospital in New York in November 2011, the study later received approval from Western IRB. Participants were invited to complete the questionnaire by visiting the study's website, www.KeloidSurvey.com. After reviewing the consent form, adult participants provided electronic consent, while parents could consent on behalf of their minor children. The questionnaire collected detailed information, including patient demographics, family history, the extent and distribution of keloid lesions, prior treatments, and responses to those treatments. To ensure data integrity, access to the survey was restricted to one entry per computer IP address. Participants had the option to skip questions that were either not applicable or that they preferred not to answer. An analysis of participants perceptions regarding the effectiveness of steroid injections has been published [1]. The dataset used for this study was accessed in May 2024.

#### Feature Description

In Supplementary Figures 1a–h, you will find the dataset and descriptive analyses covering participant demographics, age of keloid onset, ethnicity, gender distribution, Race of patients, triggering factors, lesion distribution patterns, and lesion appearance and shape.

**Gender**: The data in Supplementary Figure S1a shows that nearly twice as many female patients as male patients were diagnosed with keloids, which aligns with previous research suggesting that female sex may be a potential risk factor for keloid formation. Studies have indicated that keloid onset often occurs during puberty for both sexes, with females showing a higher prevalence, especially before age 15. This female predominance may be linked to physiological factors, including hormonal influences, and warrants further investigation into the role of sex hormones in keloidogenesis [2].

**Race and nationality**: Participants were asked to indicate their ethnic background, and Supplementary Figure S1b represents the distribution of various ethnic groups among the respondents. Although the data may not fully represent the actual ethnic distribution of keloids in the broader population, the figure shows a higher incidence of keloids among African American participants. This observation is consistent with previously published studies, which have reported a greater predisposition to keloids in this group [4, 5].

**Age**: Participants were requested to provide their current age, as well as the age at which they first noticed the development of a keloid. Supplementary Figure S1d and e respectively illustrate both the participants' ages at the time of completing the survey and the age at which their keloid initially appeared. As shown in this figure, most patients in our cohort showed keloid onset before age 20, with many cases starting as early as age 8. This is consistent with previous reports, suggesting that individuals predisposed to keloids tend to develop them early, particularly during adolescence [2, 6].

**Intralesional Triamcinolone Acetonide Injections**: Participants were asked for the total number of steroid injections they had received for their keloids, which is shown in Supplementary Figure S1f. The survey used the term steroid injection instead of ILCS, as it was presumed that participants might not be familiar with the medical terminology [8]. The survey also presumed that all patients had been treated with either TA or a similarly potent steroid. Additionally, participants were asked to recall how frequently they received these injections. The survey relied on patients' own recollections and understanding of the treatments they underwent. However, the survey did not inquire about the specific type of steroid or the dosage administered. Notably, a significant number of participants reported receiving over 10 injections, often monthly. The effectiveness of ILCS was evaluated by asking participants to select only one response to each of three multiple-choice questions, which are quoted below exactly as they appeared in the survey, and the responses are shown in Figure S2.

**Pattern of Distribution of Keloid Lesions**: Participants were requested to provide detailed information regarding the distribution of their keloid lesions across their skin. The data shows in Supplementary Figure S1f that the chest and shoulder areas are the most affected, while the ankle and forehead are the least. This distribution is consistent with other studies, suggesting that keloids are more likely to form in regions of high skin tension and frequent movement [7].

**Keloid triggering factors:** Participants were asked to share information about the factors that potentially triggered the formation of their keloids. According to the data that is shown in Supplementary Figure S1g the participants, acne was the most frequently reported trigger, with skin injury and surgery being the next most common factors. These findings are consistent with previously published data [3].

**Appearance and Shape of Keloid Lesions**: Participants were asked to describe the shape and appearance of their keloid lesions. To assist with this, an online reference guide was provided, offering categories such as flat, nodular, linear, massive, pedunculated, and superficially spreading Supplementary Figure S1h.

**Changes in keloid size over time:** As part of the survey, participants were asked to compare the current size of their keloids to their size 1, 2, 5, and 10 years ago. Their responses are summarized in Supplementary Figure S2a-e.

**Keloid size compared to 1, 2, 5 and 10 years ago**: Participants were also asked to compare the size of their keloids 1-10 years after beginning steroid treatment, with the responses providing insights into how the size of their keloids changed over this period, as summarized in Supplementary Figure S3a-d.

**Assessment of treatment response to steroid therapy:** Participants were asked to evaluate the effectiveness of steroid therapy for their keloid lesions. Their responses are summarized in Supplementary Figure S4a-c, which includes perceived outcomes (no improvement, worsening, improvement, or complete cure), changes in lesion size following treatment (i.e., final outcome of steroid injections), and classification of patients as either sensitive or resistant to steroid injections.

**Feature Engineering:** From the self-reported keloid patient dataset described above, the following variables were selected for analysis: gender, race, keloid shape, patient age, age at keloid onset, keloid history at 1, 2, 5, and 10 years prior, keloid location, keloid size changes over 1, 2, 5, and 10 years, trigger factors, and number of steroid injections administered.

After filtering the dataset to focus only on steroid therapy related data, features with significantly fewer rows were removed from the analysis to ensure robustness. Subsequently, the remaining numerical features were standardized to a range of 0 to 1 to address the disparity in value ranges and optimize model performance. Categorical features were then one-hot encoded. One-hot encoding converts categorical variables into a binary matrix representation, enabling ML algorithms to interpret the data more effectively. Each category is represented as a separate binary feature, ensuring that no ordinal relationships are assumed between categories. Following these steps, nine categories of features with different attributes remained Figure 1B on which our ML models were built.

**Handling Class Imbalance:** Class imbalance is a common issue in many ML tasks, particularly in clinical datasets where the number of instances in one class significantly outnumbers those in another. This imbalance can lead to biased model performance, where the model becomes more proficient at predicting the majority class while underperforming on the minority class. To address this, various resampling techniques can be applied to balance the class distribution, improving the model's ability to generalize to both classes. Synthetic Minority Over-sampling Technique (SMOTE) is a widely used method for addressing class imbalance. It generates synthetic examples of the minority class by interpolating between existing minority class instances. Specifically, SMOTE selects two or more similar instances in the minority class and creates a new synthetic instance along the line segment joining them. This approach helps to increase the representation of the minority class without simply duplicating the existing instances, thus providing a more diverse training set for the model. Edited Nearest Neighbors (ENN) is an under-sampling technique used to clean the dataset by removing noisy and borderline examples. ENN removes instances from the dataset that are misclassified by their k-nearest neighbors (k=3 used). This helps in eliminating potential outliers and instances that might be ambiguously labeled, leading to a more distinct class boundary.

SMOTEENN is a hybrid approach that combines the strengths of SMOTE and ENN. It first applies SMOTE to generate synthetic instances of the minority class and then applies ENN to remove noisy and borderline examples. This combination helps to not only balance the class distribution but also enhance the quality of the training set by reducing noise and improving the class boundaries. For our dataset, we evaluated several sampling strategies, and SMOTEENN delivered the best performance according to our evaluation metrics. This combination of generating synthetic instances and cleaning the dataset effectively handled the class imbalance in our keloid patient dataset, resulting in a more robust and generalizable model for predicting response to steroid therapy.

**Data Splitting and Evaluation Metrics:** To develop reliable ML models and evaluate their performance on unseen data, it is essential to split the dataset into training and testing subsets. This approach ensures that models are trained on one portion of the data (the training set) while their prediction accuracy is evaluated on a separate portion (the test set). For this purpose, the dataset was first randomly shuffled, and a 5-fold cross-validation technique was applied. This method involves dividing the dataset into five equal parts. During each iteration, one part is set aside as the test set, while the remaining four folds are used to train the model. This process is repeated five times, with each fold serving as the test set exactly once. Consequently, the benchmark results reported in the following sections are the averages of these five iterations. The predictive performance of the ML models was evaluated using several metrics. Accuracy, which measures the ratio of correctly predicted instances to the total instances, was used to provide a general measure of model performance. Precision, the ratio of true positive predictions to the total predicted positives, indicates the accuracy of positive predictions. Recall, the ratio of true positive predictions to the total actual positives, reflects the model's ability to identify all relevant instances. The F1 score, which is the harmonic mean of precision and recall, offers a single metric that balances both concerns. Additionally, the F0.5 score was utilized to evaluate performance, as it places greater emphasis on precision. This choice was made because false positives are particularly costly in our context. A false positive result would incorrectly indicate the need for a steroid injection, leading to un- necessary treatment for the patient. The ROC-AUC (Receiver Operating Characteristic Area Under the Curve) metric was employed to measure the model's ability to distinguish between classes, with higher values indicating better performance. Lastly, the PR-AUC (Precision-Recall Area Under the Curve) metric was used to assess the trade-off between precision and recall across different threshold settings, which is particularly useful for imbalanced datasets. By using these metrics, we ensure a comprehensive evaluation of our models, capturing their performance across various critical aspects of the task at hand, and understanding the strengths and weaknesses of each model, guiding further improvements and optimizations.

**Machine Learning (ML) models:** To predict the response to steroid therapy in keloid patients, we applied a range of ML models (Table 1). Given the complexity and variability in keloid lesion formation, it was essential to explore different models to capture non-linear and intricate relationships within the data. Each model offers unique advantages in terms of classification capability and generalizability. By systematically comparing these models, we aimed to identify the one best suited for this predictive task. Below, we provide a brief overview of each model and its implementation.

*Random Forest (RF)* - RF, an ensemble learning algorithm, excels in regression applications [9]. RF integrates bagging and bootstrap aggregating techniques. During bagging, RF generates multiple decision trees simultaneously, each trained on a randomly selected subset of the dataset, allowing for replacement. This strategy enhances tree diversity, mitigates overfitting, and improves generalization. Bootstrap aggregating involves sampling with re- placement from the original dataset to create multiple subsets for training. This approach ensures that each decision tree in the RF ensemble encounters different data instances, bolstering the model's robustness and decreasing variance. During training, each tree in the RF ensemble independently predicts the target variable. For regression tasks, RF combines all tree predictions to form a final prediction, typically by averaging their outputs. This ensemble methodology often results in better predictive performance compared to single decision trees. A significant benefit of RF is its ability to handle high- dimensional datasets with numerous features, as it automatically selects a subset of features for each split, thus reducing overfitting risk and enhancing computational efficiency.

*Gradient Boosting (GB) Trees* - GB is another ensemble technique leveraging decision trees for predictions [10]. Unlike RF, GB uses shallow decision trees, or weak learners, and combines their outputs during training. GB builds trees sequentially, with each new tree aiming to correct errors made by its predecessors. By iteratively refining these errors, GB constructs a strong predictive model over time.

A key strength of GB is its ability to uncover complex relationships in the data, as the model continuously learns from its mistakes and improves predictions. Additionally, GB is less prone to overfitting compared to deep decision trees, making it suitable for various regression tasks. However, GB typically demands more computational resources and fine- tuning than Random Forest, given its sequential learning process. Despite this, GB often delivers superior predictive performance, particularly on structured datasets with intricate patterns [11].

*Neural Network (NN)* - NNs, a subset of ML, underpin deep learning techniques. Inspired by the human brain's biological structure, NNs consist of interconnected neurons that communicate. These networks usually have an input layer, one or more hidden layers, and an output layer. During training, NNs undergo an iterative process where initial random weights are adjusted via backpropagation until the model minimizes the loss function, converging to a global minimum. By continuously fine-tuning weights based on observed errors, NNs can effectively learn complex data patterns and relationships.

NNs are highly valued for their capacity to handle large and varied datasets and to learn hierarchical data representations. They are particularly effective in image and speech recognition, natural language processing, and other tasks. However, training deep neural networks can be computationally demanding and may require substantial labeled data. Nonetheless, NNs lead many state-of-the-art ML applications, driving advancements across numerous fields.

*XGBoost* **-** XGBoost, or Extreme Gradient Boosting, is an advanced variant of Gradient Boosting algorithms, incorporating regularization techniques like L1 and L2 for better generalization [12]. Unlike traditional gradientboosted trees, XGBoost offers superior speed, high performance, and the ability to conduct parallel tree boosting. Its efficiency makes XGBoost suitable for large-scale datasets and computationally intensive tasks. XGBoost is popular for its robustness and versatility across various ML applications.

In our study, XGBoost is also used for feature importance analysis and SHAP (SHap- ley Additive exPlanations) analysis. These techniques help practitioners understand the relative importance of input features and the model's decision-making process. This dual functionality enhances XGBoost's interpretability, making it a valuable tool for both predictive modeling and interpretability in ML tasks.

*Support Vector Classifier (SVC)* - Support Vector Machines (SVMs) are a class of ML algorithms used for both regression and classification tasks. For the classification task, SVMs predict by finding a hyperplane that separates data points of different categories by maximizing the distance, called the margin, between the nearest data points and the decision boundary [13]. For the regression task, SVMs provide a decision boundary at a distance from the original hyper- plane such that the data points closest to the hyperplane, called support vectors, are within that boundary line.

In this work, we utilized the SVC, a SVM variant for classification. SVC finds the optimal hyperplane by focusing on support vectors, improving generalization to new data. Kernel functions allow SVC to handle non-linear relationships, making it versatile for various classification problems. SVC is effective in high-dimensional spaces and robust to overfitting, particularly when features outnumber samples.

*Logistic Regression (LR)* **-** LR is a widely-used statistical method for binary classification tasks [10]. LR models the probability of a binary outcome by fitting a logistic function to the input features. This approach transforms the linear combination of features into a probability score between 0 and 1. The model's coefficients are estimated using maximum likelihood estimation, and the resulting probabilities are used to classify observations into one of two categories. One of the strengths of LR is its interpretability, as the coefficients can provide insights into the influence of individual features on the outcome. Despite its simplicity, LR can per- form well on a variety of datasets, particularly when the relationship between the features and the outcome is approximately linear. However, LR may struggle with complex, non-linear relationships and typically requires feature scaling and careful handling of multicollinearity among predictors.

*CatBoost* **-** CatBoost is a gradient boosting algorithm developed by Yandex, designed specifically to handle categorical features effectively [14]. Unlike many other algorithms that require pre-processing of categorical data into numerical form, CatBoost can directly process categorical variables, preserving their structure and relationships. This capability often leads to better performance and easier model deployment.

CatBoost employs ordered boosting, a variant of gradient boosting that reduces overfit- ting and variance by processing data in a permutation-driven manner. This approach helps in maintaining the robustness of the model. CatBoost is known for its fast training times, ease of use, and strong performance on a wide range of datasets, making it a popular choice in the ML community.

*Bagging* **-** Bagging, short for Bootstrap Aggregating, is an ensemble learning technique that enhances the stability and accuracy of ML algorithms [15]. Bagging involves creating multiple versions of a predictor by training each version on a different bootstrap sample from the original dataset. The final prediction is obtained by averaging (for regression) or voting (for classification) across all predictors.

This method reduces variance and helps prevent overfitting, especially for high-variance models like decision trees. By training multiple models on different subsets of the data, Bagging produces a more robust and generalizable predictive model. One of the notable advantages of Bagging is its simplicity and ability to improve the performance of many base models without requiring significant modifications.

*AdaBoost* **-** AdaBoost, short for Adaptive Boosting, is an ensemble technique that combines the out- puts of weak learners to create a strong classifier [16]. Unlike Bagging, which trains multiple models independently, AdaBoost builds models sequentially. Each new model focuses on the instances that the previous models misclassified, adjusting their weights to improve performance on these hard-to-classify examples.

AdaBoost works by assigning higher weights to incorrectly classified instances and lower weights to correctly classified ones. This iterative process continues until a predetermined number of models are trained or no further improvement is observed. AdaBoost is known for its simplicity, effectiveness, and ability to reduce both bias and variance. However, it can be sensitive to noisy data and outliers, which may affect its performance if not properly managed.

For our keloid patient dataset, we implemented each of these models and evaluated their performance using comprehensive metrics. This thorough approach allowed us to identify the most effective model for predicting response to steroid therapy, after accounting for the specific challenges posed by our data.

*Hyperparameter Optimization* - Hyperparameter optimization (Supplementary Table S1) is crucial in ML to finetune models for optimal performance. For Random Forest, the hyperparameters were tuned including n estimators, max depth, min samples split, min samples leaf, max features, and bootstrap. These parameters control the number of trees in the forest, the maximum depth of each tree, the minimum number of samples required to split an internal node, the minimum number of samples required to be at a leaf node, the number of features to consider when looking for the best split, and whether bootstrap samples are used when building trees.

SVC optimization focused on parameters such as C, gamma, and kernel. The C parameter balances the trade-off between achieving a low training error and minimizing model complexity, gamma defines the influence of a single training example and affects the flexibility of the decision boundary, and kernel specifies the kernel type used in the algorithm (linear, polynomial, sigmoid, or radial basis function), impacting the decision function shape and model flexibility.

Logistic Regression's hyperparameters, including C, solver, penalty, and max-iter, were optimized to enhance model regularization, optimization method, regularization type, and convergence criteria, respectively. These parameters collectively govern model fitting and performance optimization, crucial for achieving robust logistic regression models.

Gradient Boosting tuned parameters such as n estimators, learning rate, max depth, subsample, and min samples split. These parameters control the number of boosting stages, the shrinkage of the contribution of each tree, the maximum depth of each tree, the fraction of samples used for fitting each tree, and the minimum number of samples required to split an internal node, collectively enhancing model fitting and generalization through boosting.

Neural Network optimization involved parameters like hidden layer sizes, activation, solver, alpha, learning rate, and max-iter. These parameters define the network architecture, activation functions for hidden layers, weight optimization strategy, regularization strength, learning rate schedule, and maximum number of iterations, influencing the neural network's learning dynamics and performance.

XGBoost parameters, including n estimators, learning rate, max depth, subsample, and colsample bytree, were tuned to optimize the number of boosting rounds, control overfitting by learning rate adjustment, limit tree depth and instance subsampling, and select the fraction of features used per tree, enhancing prediction accuracy and efficiency through boosting.

CatBoost optimized iterations, learning rate, depth, and l2 leaf reg parameters. These parameters control the number of boosting iterations, step size during optimization, tree depth, and leaf regularization, respectively, optimizing boosting process characteristics to improve model training and performance.

Bagging's hyperparameters, including base estimator max depth and n estimators, were tuned to restrict base model depth and determine the number of base estimators, influencing ensemble complexity and robustness for improved prediction accuracy.

AdaBoost's parameters, n estimators and learning rate, were optimized to define the maximum number of boosting stages and weight the contribution of each classifier, respectively, governing boosting round execution and impact on model learning and prediction.

The optimization details, along with the best-performing hyperparameter values, are summarized in Supplementary Table S1. This table offers a comprehensive overview of the key hyperparameters for each model and their optimal values.

**Feature Importance Analysis:** In this study, two complementary methods—SHAP (Shapley Additive Explanations) and Permutation Importance—were used to evaluate feature importance and interpret the model's predictions. These methods offer distinct but valuable perspectives on the relative influence of input features.

*SHAP (Shapley Additive Explanations)* - SHAP, grounded in cooperative game theory, was employed for comprehensive feature importance analysis [17]. SHAP assigns an importance value to each feature by calculating its contribution to the model's predictions, based on all possible combinations of features. This makes it particularly useful for capturing interactions between features, which is crucial in models where features collectively influence predictions. The framework ensures that the sum of the feature importance values equals the model's total output, providing both mathematically rigorous and interpretable results.

SHAP allows for global interpretability, identifying the most influential features across the entire dataset, and local interpretability, examining the specific impact of features on individual predictions. This dual capability is essential in understanding how different combinations of clinical features contribute to model predictions.

In this study, SHAP revealed the importance of key clinical features such as gender, age, and growth history of keloid lesions, not only as individual predictors, but also in their interactions with other features. This deeper understanding of the model's internal mechanics enabled us to refine our model, focusing on the most critical features and ensuring that the predictions were consistent with clinical knowledge. Using SHAP, we were able to maintain a transparent and interpretable model, aligning predictive insights with clinical expertise and strengthening the credibility of our findings.

*Permutation Importance* **-** To complement SHAP, we employed permutation importance to provide a more global measure of feature importance. Permutation importance evaluates how much each feature contributes to the overall model performance by randomly shuffling its values and observing the resulting drop in accuracy [9]. This method provides a straightforward assessment of how the model depends on each feature, independent of interactions.

The strength of permutation importance lies in its simplicity, offering a clear, global view of feature influence. Unlike SHAP, which excels at capturing feature interactions, permutation importance focuses on each feature's individual impact. Together, these methods offer a comprehensive perspective on feature importance, balancing the nuanced interactions identified by SHAP with the direct, global influence revealed by permutation importance. Permutation importance was calculated by measuring the model's accuracy before and after shuffling each feature. A significant drop in accuracy indicated the feature's importance in making predictions. Like SHAP, permutation importance was also aggregated at the group level to evaluate clinically relevant feature sets.

By combining SHAP and permutation importance, we gain a more holistic understanding of feature importance. SHAP provides the nuance of feature interactions and individualized predictions, while permutation importance offers a straightforward and intuitive ranking of feature influence on global model performance. Together, these methods ensure that we not only understand which features the model relies on but also how these features interact to drive clinical predictions.

**The data and code associated with this study will be made available upon a written request.**

***Supplementary References:***


1. Tirgan, M. *Worsening of keloids after intralesional injections*. in *Journal of the American Academy of Dermatology*, 2013. **68**: p. AB68.
2. Chikage, N., Y. Hayasaka, and R. Ogawa, *Sex Differences in Keloidogenesis: An Analysis of 1659 Keloid Patients in Japan.* Dermatology and Therapy, 2019. **9**: p. 747 - 754.
3. Wen-sheng, L., et al., *Clinical and epidemiological analysis of keloids in Chinese patients.* Archives of Dermatological Research, 2015. **307**: p. 109-114.
4. Swenson, A., et al., *Natural History of Keloids: A Sociodemographic Analysis Using Structured and Unstructured Data.* Dermatology and Therapy, 2024. **14**(1): p. 131-149.
5. Tot, L., et al., *Textbook on scar management: state of the art management and emerging technologies.* 2020.
6. Ogawa, R., et al., *The relationship between skin stretching/contraction and pathologic scarring: the important role of mechanical forces in keloid generation.* Wound Repair and Regeneration, 2012. **20**(2): p. 149--157.
7. McGinty, S. and W.J. Siddiqui, *Keloid.* 2018.
8. Tirgan, M., *Intralesional triamcinolone acetonide in the treatment of keloid lesions-can the treatment be harmful to some patients? results of an online survey.* Int J Keloid Res, 2017. **1**: p. 21-28.
9. Breiman, L.
10. Friedman, J., T. Hastie, and R. Tibshirani, *Additive logistic regression: a statistical view of boosting (with discussion and a rejoinder by the authors).* The annals of statistics, 2000. **28**(2): p. 337--407.
11. Hinton, G.E., S. Osindero, and Y.-W. Teh, *A fast learning algorithm for deep belief nets.* Neural computation, 2006. **18**(7): p. 1527-1554.
12. Chen, T. and G. Carlos, *"XGBoost: A Scalable Tree Boosting System.".* Proceedings of the 22nd ACM SIGKDD International Conference on Knowledge Discovery and Data Mining, 2016: p. 785–94.
13. Cortes, C. and V. Vapnik.
14. Prokhorenkova, L., et al., *CatBoost: unbiased boosting with categorical features.* Advances in neural information processing systems, 2018. **31**.
15. Breiman, L., *Bagging predictors.* Machine learning, 1996. **24**: p. 123--140.
16. Freund, Y. and R.E. Schapire, *A decision-theoretic generalization of on-line learning and an application to boosting.* Journal of computer and system sciences, 1997. **55**(1): p. 119--139.
17. Lundberg, S.M. and S.-I. Lee, *A unified approach to interpreting model predictions.* Advances in neural information processing systems, 2017. **30**.

**SUPPLEMENTARY FIGURE LEGENDS**:

**Figure S1:** Distribution of patients based on **(a)** gender, **(b)** race or nationality, **(c)** age of patient, **(d)** age of keloid onset, **(e)** number of steroid injections, **(f)** keloid location, **(g)** keloid trigger, and **(h)** keloid shape. Representative images of different keloid morphologies referred to in panel h, namely (I) nodular, (II) linear, (III) flat, (IV) massive, (V) superficially spreading, and (VI) pedunculated can be found at: Tirgan, M. H. Types of keloid lesions, Keloid 212. Retrieved April 29, 2025, from https://www.keloid212.com/types-of-keloid-lesions/.

**Figure S2:** Distributions of patients based on the number of keloids at different time points. **(a)** present day, **(b)** 1 year ago, **(c)** 2 years ago, **(d)** 5 years ago, and **(e)** 10 years ago.

**Figure S3**: Distribution of patients in the dataset based on changes in the size of their keloids over time, when compared to **(a)** 1 year ago, **(b)** 2 years ago, **(c)** 5 years ago, and **(d)** 10 years ago.

**Figure S4:** Self-reported patient survey results on the effectiveness of steroid therapy. **(a)** Patient responses categorized by perceived benefit following steroid injections: “No improvement”, “Condition worsened”, “Improved”, and “Fully cured”. **(b)** Outcomes following steroid treatment: “Improved but returned to original size”, “Became larger”, “Improved and remained smaller”, and “Completely resolved”. **(c)** Classification of patients based on response to steroid treatment as “Resistant” or “Sensitive” to steroid injections.

**Supplementary Table S1: Hyperparameter Optimization Details with Best Parameters**

| ML Model | Hyperparameter | Search Space | Best Parameters |
|---|---|---|---|
| Random Forest | n_estimators | 100, 200, 300 | 200 |
| | max_depth | None, 10, 20, 30 | 20 |
| | min_samples_split | 2, 5, 10 | 2 |
| | min_samples_leaf | 1, 2, 4 | 1 |
| | max features | 'auto', 'sqrt', 'log2' | 'sqrt' |
| | bootstrap | True, False | False |
| SVC | C | 0.1, 1, 10, 100 | 10 |
| | gamma | 1, 0.1, 0.01, 0.001 | 1 |
| | kernel | 'linear', 'rbf', 'poly', 'sigmoid' | 'linear' |
| Logistic Regression | C | 0.1, 1, 10, 100 | 10 |
| | solver | 'newton-cg', 'lbfgs', 'liblinear' | 'newton-cg' |
| | penalty | 'l2' | 'l2' |
| | max_iter | 100, 200, 500 | 100 |
| Gradient Boosting | n_estimators | 100, 200, 300 | 300 |
| | learning_rate | 0.01, 0.1, 0.5, 1.0 | 0.1 |
| | max_depth | 3, 4, 5, 6 | 6 |
| | subsample | 0.6, 0.8, 1.0 | 0.6 |
| | min_samples_split | 2, 5, 10 | 2 |
| Neural Network | hidden_layer_sizes | (64, 128, 128), (128, 128), (128, 256, 256) | (128, 256, 256) |
| | activation | 'relu', 'tanh' | 'relu' |
| | solver | 'adam', 'sgd' | 'adam' |
| | alpha | 0.0001, 0.001, 0.01 | 0.0001 |
| | learning_rate | 'constant', 'adaptive' | 'constant' |
| | max_iter | 20000 | 20000 |
| xgb | n_estimators | 100, 200, 300 | 100 |
| | learning_rate | 0.01, 0.1, 0.5 | 0.5 |
| | max_depth | 3, 4, 5, 6 | 6 |
| | subsample | 0.6, 0.8, 1.0 | 1.0 |
| | colsample_bytree | 0.6, 0.8, 1.0 | 1.0 |
| catboost | iterations | 100, 200, 300 | 300 |
| | learning_rate | 0.01, 0.1, 0.5 | 0.5 |
| | depth | 3, 4, 5, 6 | 5 |
| | l2_leaf_reg | 3, 5, 7 | 7 |
| bagging | base_estimator max_depth | None, 10, 20 | None |
| | n_estimators | 50, 100, 200 | 100 |
| adaboost | n_estimators | 50, 100, 200 | 200 |
| | learning_rate | 0.01, 0.1, 1.0 | 1.0 |
